# Value of chest Computed Tomography imaging radiomics features in predicting breast density classification

**DOI:** 10.1101/2025.01.05.25320024

**Authors:** Weipeng Zhou, Qi Yang, Huimao Zhang

## Abstract

**Objective:** To explore the correlation between the radiomic characteristics derived from chest CT scan imaging and breast density classification, to construct an imaging radiomics model that can automatically achieve CT breast density classification, and to evaluate its diagnostic efficacy, so as to provide a valuable reference for breast density AI classification through chest CT scanning images.

**Methods:** 393 patients who had negative results of breast ultrasound and mammography in our hospital and had undergone CT scanning of the chest within one year were collected for retrospective analysis, and 330 patients who had consistent results of breast density classification judged by two radiologists based on mammography images were used for the construction of imaging radiomics prediction models, which involved mapping of three-dimensional regions of interest (ROI) of the breast, extraction of imaging radiomics features and dimensionality reduction, screening of dominant radiomics features, establishment of multiple four-classification prediction models, and evaluation the effectiveness of each model.

**Results:** A U-net neural network segmentation model was trained to automatically delineate breast ROI, extract 1427 radiomics features, and screen out 28 dominant features closely related to breast density for AI automatic classification of breast density. Among the four types of four-classification prediction models constructed based on classifier including ➀ Xgboost Classifier, ➁ One Vs Rest Classifier(Logistic Regression in the One Vs Rest framework), ➂ Gradient Boosting, ➃ Random Forest Classifier, Xgboost four-classification prediction model has the best prediction performance, and after its parameter tuning, the classification accuracy for the test set reaches 0.866, and the area beneath the curve (AUC) derived from the receiver operating characteristic curves (ROC) of the four categories of classification labels are 1.00, 0.93, 0.93,and 0.99, respectively, and the AUC of the micro-averaged ROC of each classification label is 0.97, and the AUC of the macro-averaged ROC is 0.96, indicating the best prediction performance.

**Conclusion:** Chest CT plain images can provide breast density classification, a valuable information that reveal significant insights pertaining to breast cancer risk, and can be used to automatically achieve breast density four-classification through imaging radiomics model, which lays the foundation for precise and individualized breast screening programs. Introducing AI methods for automatic breast density classification can help clinical diagnosis.

**Author summary:** Weipeng Zhou is the scheme designer of this paper, and is responsible for collecting relevant data, conducting data analysis and modeling, writing and revising the manuscript. Huimao Zhang is the corresponding author of the paper, responsible for scheme design and paper revision. This paper is funded by the fund project hosted by Huimao Zhang. Jianhua Liu is the co-author of this paper, responsible for data analysis.

## 1 Introduction

In the 21st century, there has been a notable rise in both the rates of breast cancer diagnosis and the associated mortality. The incidence of breast cancer among women has emerged as the most prevalent form of cancer globally.

The number of breast cancer incidence and deaths is also on the rise in china, and is expected to increase by 36.3% and 54.0% respectively by 2030 [1], imposing a heavy disease and economic burden on women. Therefore, it is important to accurately risk-stratify the occurrence of breast cancer and select appropriate imaging screening programmes accordingly.

The density of breast tissue indicates the ratio of glandular tissue to adipose tissue within the breast. According to the fifth edition of the BI-RADS, there are four distinct categories into which breast density can be classified [2]. A higher level of breast density is significantly associated with an increased likelihood of developing breast cancer, which can contribute to the development of breast tumors [3]. In countries like the United States, there are existing legislations that require informing patients who undergo mammography about their breast density [4]. Supplementary breast cancer screening programs, such as magnetic resonance imaging, have been recommended for women with high breast density, which may increase the cancer detection rate in these women [3–6].

Although previous studies have shown that beginning annual mammography screenings at the age of 40 may yield significant benefits [7], the adherence of Chinese women to specialized breast cancer screening is still far from sufficient. Conversely, with the continuous improvement in people’s understanding of screening for diseases like lung cancer, the volume of CT examinations is increasingly surging.The feasibility of breast density assessment on CT has previously been demonstrated by Salvatore et al. in a study conducted in 2014 [8].

It is not uncommon for CT to be used to provide additional information beyond the primary indication, for example, chest CT for lung screening has also been used to provide a lot of additional value in cardiovascular disease [9, 10]. Therefore, for many women who have undergone chest CT scans but not mammography, valuable information about breast cancer risk can similarly be provided to these women if breast density can be reliably determined from CT images [7, 8, 11].

This study aims to evaluate the concordance between chest CT and mammography in breast density assessment through the interpretation of images by radiologists, and to realize automatic discrimination of breast density types through chest CT using radiomics methods, so as to provide this risk related information of breast cancer and offering new insights for breast cancer screening.

## 2 Data and Methods

### 2.1 Research participant

Retrospectively, we analyzed the medical records of breast-healthy patients who underwent mammography and breast ultrasound imaging in our hospital from January 2021 to December 2022 with negative results and underwent chest CT scan within 12 months of the mammography. Inclusion criteria: (1) the results of mammography and ultrasound examination were negative; (2) chest CT scan was performed in our hospital within 1 year. Exclusion criteria: (1) patients were found to have breast lesions in any examination; (2) incomplete images; (3) poor image quality or incompatibility with machine learning models. Approval for this study was granted by the Ethics Committee at the hospital.

This study was approved by the Ethical Committee of the First Hospital of Jilin University and exempted from the requirement for informed consent. The retrospective study accessed data on October 10, 2024, and during or after the data collection period, it was not possible to access identifiable information of individual participants.

### 2.2 Collection of data and establishment of models

Mammography images and non-contrast chest CT scan images were collected, the images were classified into four types according to breast density by two radiologists using a blind method respectively. Patients with consistent results in terms of breast density classification judged by the two radiologists based on the mammography images were included in the dataset.

After completing the layer-by-layer identifying and c of the four ROIs (left breast, right breast, left axilla, and right breast) on the sequence images of chest CT without contrast, feature extraction, feature screening and dimensionality reduction were carried out to obtain the dominant features, and the dataset of the dominant features was used to construct various four-classification prediction models, evaluate the prediction performance of each model, and compare them to obtain the optimal model.

#### 2.2.1 Image acquisition

The CT machine models used in this study were PHILIPS iCT64/256. The scanning parameters were set as follows: Detector configuration: number of layers: 64, detector width: 0.625 mm (single-layer detector width), total detector width: 40 mm. The tube operated at a voltage of 120 kVp, with the current being adjusted automatically. Pitch was set to 5. Image reconstruction matrix: standard matrix was 521 × 521 pixels, reconstruction layer thickness was 1-1.5 mm. After all images were scanned, they were uniformly processed into thin-layer CT images with a thickness of 1mm and archived in DICOM format for storage in the PACS system.

The mammography machine used in this study was Hologic Lorad M-I. The parameters for the scanning process were established as follows: 2D Mammography. Tube voltage (kV): 22 kV to 35 kV, typical 28 KV. Tube current (mA): 10 mA to 100 mA, typical 50 mA. Exposure time (sec): 0.1 s to 10 s, typical 0.5 s. Focus size: 0.1 mm for microfocus, 0.3 mm for conventional focus. Image size: Maximum film size: 18 × 24 cm.

#### 2.2.2 Analyzing images and extracting features

##### 2.2.2.1 Image analysis

According to the BI-RADS [2], each patient’s mammogram or chest CT plain scan image was classified into one of the following four types using a visual rough quartile evaluation. Category A-The breast is almost entirely composed of fat; B-There is scattered fibroglandular in the breast; C-The breast is inhomogeneously dense; D-The breast density is very high.

Weighted Kappa Statistic was used to evaluate the agreement in breast density assessment between two different examinations for each radiologist, as well as the interobserver agreement for each examination individually.

Patients with concordant four-category classification results of breast density assessed by two radiologists based on mammography images (considered as criteria) were included in the dataset for the development of the four-category classification model of breast density.

##### 2.2.2.2 Breast ROI segmentation

In this study, we used the self-developed RIAS platform to complete the ROI delineation work, and completed the layer-by-layer delineation and verification of the four ROIs of the bilateral breasts and axillae on the serial images of the chest CT scans, ROI delineation diagram is shown in Fig.1.

**Fig 1.**
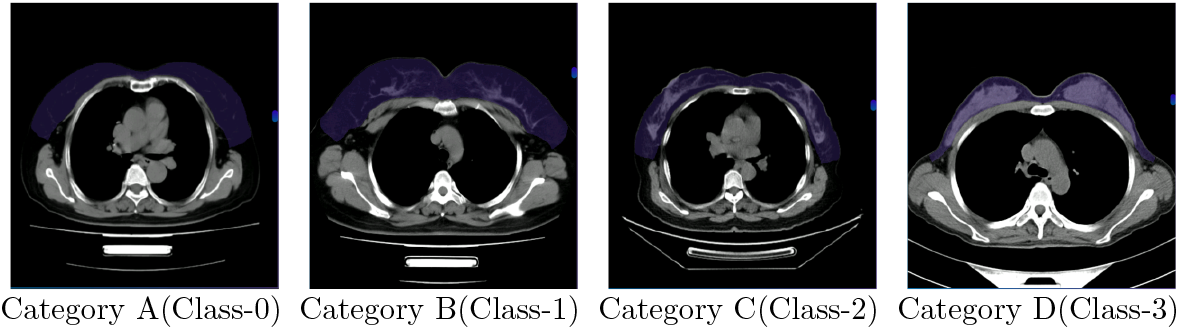
Breast ROI delineation diagram

Due to the unclear boundaries between the breast and surrounding structure on CT images, this study, by combined imaging interpretation experience and the anatomical theory of the superficial fascial system of the breast [12], had established for the first time a delineation criteria for breast ROIs. For a unilateral breast, the upper boundary is defined by the level where the first rib extends to the midclavicular line on a CT image, and the lower boundary is at the level where the sixth rib extends to the midclavicular line. The inner boundary is marked by the body’s midline, and the outer boundary is defined by the posterior edge of the axilla. This delineation criteria ensures good consistency and reproducibility in the selection of ROIs.

After manually delineating all the enrolled chest CT images layer by layer, we established an automatic segmentation model based on the U-net deep learning algorithm. The segmentation performance of the model was evaluated using the Dice Similarity Coefficient.

##### 2.2.2.3 Feature extraction and dimensionality reduction from breast ROI region

Morphological features, texture features, first-order statistical features, filtered features and other radiomics features from the entire breast ROI region were extracted using Pyradiomics software(http://pyradiomics.html). The features were first screened initially by calculating the intraclass correlation coefficient (ICC). Then, screened the features by t-test. Finally, further refine the feature selection using the LassoCV regression model.

###### Feature Testing

A univariate model (fitted model of the data of individual feature variable to the classification outcome) was fitted using logistic regression, and the summary information from the model was analyzed to determine whether the features selected through the above screening have a significant impact on the classification results (*p* < 0.05).

#### 2.2.3 Development and assessment of the radiomic model’s efficacy

Python (version3.6) was used to up-sample the four-class classification data to address the issue of imbalance in the data. Four four-classification machine learning models were established. The assessment of model efficacy for every category was conducted utilizing ROC curves along with Decision Curve Analysis(DCA). The accuracy of the models’ predictions for each category was assessed through the use of calibration curves. Moreover, both macro-level and micro-level averaging techniques were employed to assess the comprehensive effectiveness of the models.

### 2.3 Standards for categorizing breast density

The 5th edition of BI-RADS introduces specific percentile quartile standards for evaluating breast conditions. The assessment of breast density was based on a general visual evaluation utilizing a quartile system rather than relying on specific density measurements [2, 13]. Mammography has been widely used for early breast cancer screening in asymptomatic women for 40 years and is considered to be the gold standard for imaging [14]. Therefore, in this study, the results of the breast density classification determined from mammographic images were considered as the standard results for the patient’s breast density type.

### 2.4 Statistical analysis

The analysis of statistical data was conducted utilizing RStudio(version 4.4). For the measurements (data of each characteristic), the data were divided into 4 groups for ANOVA according to the 4 classification labels(where **class 0** is link to Category A, **class 1** is associated with Category B, **class 2** is associated with Category C, **class 3** is associated with Category D,) of breast density. *P* < 0.05 the statistical significance of 0.05 was recognized. Assessments of normality were conducted for each set of data.

Should the data conform to the principles of normal distribution and the consistency of variances, they were expressed as the mean ± standard deviation (m*±*sd). Otherwise, they were described using the median [interquartile range, IQR]. The consistency of breast density assessment results by each radiologist for the two different examinations, as well as the inter-observer consistency for each examination, was evaluated using the weighted Kappa statistic (Weighted Kappa Statistic, WKS). The stability of radiomics characteristics was assessed through the use of intraclass correlation coefficients (ICCs). An ICC of 0.75 or above was deemed acceptable. The evaluation of radiomics models was conducted through various performance metrics, including ROC graphs, AUC graph, and decision curve analysis (DCA), precision-recall (PR), accuracy, specificity, sensitivity, F1-score, positive predictive rate (PPR), negative predictive rate (NPR), micro-average, and macro-average.

## 3 Results

### 3.1 Statistical analysis

Based on the defined criteria for inclusion and exclusion, 330 women with healthy breasts, ranging in age from 23 to 79 years were included. The four classifications of breast density: 26 cases (7.88%) with class 0 (fatty type), 133 cases (40.30%) with class 1 (fibroglandular type), 149 cases (45.15%) with class 2 (unevenly dense type), and 22 cases (6.67%) with class 3 (extremely dense type). An analysis of variance (ANOVA) was performed, with *P* < 0.01. The statistical table of data for the four categories is shown in Table 1.

**Table 1.** The statistical data related to imaging feature across the four distinct categories.

| Feature name | Class 0<br>(n=26) | Class 1<br>(n=133) | Class 2<br>(n=149) | Class 3<br>(n=22) | Statistical F | P-value |
| --- | --- | --- | --- | --- | --- | --- |
| oriSlice023938 | 36[26.30] | 327[33.30] | 322[30.20] | 325[30.30] | 3.32<sup>a</sup> | 0.025* |
| oriFlatness005507 | 0.27±0.03 | 0.27±0.03 | 0.26±0.03 | 0.26±0.02 | 11.8 | 0.0007*** |
| oriLength001928 | 18±12.20 | 185±13.70 | 188±12.70 | 193±14.70 | 7.91 | 0.0054** |
| expEntropy015865 | 1[1.44] | 1[1.58] | -3.2e-16[1] | -3.2e-16[1] | 3.73<sup>b</sup> | 0.016* |
| logMean008652 | -22.3[7.99] | -25.1[13.70] | -28[15.80] | -24.1[12.80] | 6.04<sup>a</sup> | 0.001*** |
| logImc1004067 | -0.35[0.04] | -0.34[0.05] | -0.33[0.05] | -0.32[0.03] | 9.3<sup>a</sup> | 3.3e-05*** |
| logMCC003552 | 0.85[0.07] | 0.85[0.07] | 0.84[0.06] | 0.83[0.03] | 4.45<sup>b</sup> | 0.007** |
| logEmphasis026479 | 1.66e+09[4.06e+09] | 1.59e+09[2.74e+09] | 7.48e+08[1.13e+09] | 9.32e+08[7.37e+08] | 8.11<sup>b</sup> | 0.000*** |
| logSkewness047910 | 12.3[2.78] | 8.11[4.62] | 3.63[2.57] | 2.38[0.98] | 142<sup>b</sup> | 5.19e-29*** |
| logCorrelatio027572 | 0.54[0.06] | 0.59[0.11] | 0.73[0.11] | 0.76[0.09] | 83.6<sup>a</sup> | 2.08e-22*** |
| logIdmn073959 | 0.99[0.001] | 0.99[0.01] | 0.99[0.01] | 0.98[0.01] | 165<sup>b</sup> | 9.23e-35*** |
| oriPercentile002120 | -81[10.50] | -74[23.00] | -32[56.00] | -2.5[30.50] | 129<sup>b</sup> | 2.95e-29*** |
| oriCorrelatio028941 | 0.54[0.06] | 0.62[0.13] | 0.75[0.09] | 0.81[0.06] | 115<sup>b</sup> | 3.35e-26*** |
| oriMCC149813 | 0.59[0.07] | 0.68[0.11] | 0.78[0.08] | 0.83[0.05] | 112<sup>b</sup> | 4.72e-26*** |
| squDeviation002093 | 0.75[0.59] | 0.98[0.37] | 1.24[0.59] | 1.27[0.49] | 9.11<sup>b</sup> | 4.34e-05*** |
| squIdm007157 | 1[0.00] | 1[1.51e-06] | 1[0.00] | 1[0.00] | 5.01<sup>b</sup> | 0.003** |
| squNormalizd032501 | 0.5[0.65] | 0.5[0.68] | 0.695[0.67] | 1[0.50] | 5.06<sup>a</sup> | 0.003** |
| squSkewness019985 | 4.42[1.80] | 4.3[1.40] | 2.84[1.70] | 2.01[0.83] | 45.5<sup>a</sup> | 8.97e-16*** |
| wavCorrelati000177 | 0.47[0.03] | 0.46[0.04] | 0.45[0.04] | 0.43[0.03] | 10.8<sup>a</sup> | 7.05e-06*** |
| wavyNormald035628 | 0.04[0.006] | 0.04[0.005] | 0.03[0.004] | 0.03[0.003] | 5.62<sup>b</sup> | 0.002** |
| waveContrast030003 | 0.03[0.03] | 0.04[0.04] | 0.05[0.04] | 0.04[0.03] | 8.63<sup>b</sup> | 6.55e-05*** |
| waveSkewne017662 | -5.92[5.17] | -5.19[4.57] | -4.8[4.06] | -4.43[2.51] | 5.68<sup>b</sup> | 0.001*** |
| waveCorrelat010819 | 0.14[0.05] | 0.14[0.07] | 0.17[0.07] | 0.17[0.04] | 1.46<sup>a</sup> | 0.235 |
| wavekewness030251 | -0.05[0.25] | -0.11[0.37] | -0.19[0.55] | -0.28[0.75] | 3.13<sup>a</sup> | 0.031* |
| wavBusyness010195 | 9356[11006] | 7405[10686] | 6268[7554] | 4145[6068] | 4.13<sup>b</sup> | 0.009** |
| wavntropy000570 | 4.63±0.14 | 4.73±0.17 | 4.95±0.17 | 5.08±0.20 | 190.0 | 1.99e-34*** |
| wavformity010646 | 7117[1761] | 7187[1763] | 8003[1867] | 9206[1784] | 10.20<sup>a</sup> | 1.31e-05*** |
| wavtropy012349 | 6.53±0.25 | 6.67±0.20 | 6.78±0.22 | 6.86±0.18 | 48.80 | 1.6e-11*** |
| age | 56.6±9.39 | 52.3±8.46 | 47.3±7.15 | 43.3±7.9 | 62.10 | 4.96e-14*** |

**Remarks:** a: non-normal distribution, b: unequal variances; \* denotes $P < 0.05$ , \*\* denotes $P < 0.01$ , \*\*\* denotes $P < 0.001$

### 3.2 Extraction of features from radiomics and the development of predictive models

#### 3.2.1 Feature extraction

From the CT plain scan images delineated, 1427 image features of the breast region were extracted. Through inter-observer and intra-observer consistency analysis (ICCs), 1338 features with ICC≥0.75 were selected, while 89 features with ICC<0.75 were screened out. After t-test preliminary screening, 757 features (feature variables with classification effects) were obtained. Further feature selection was performed using the LASSO algorithm.

As the hyperparameter alphas changed, the mean squared error (MSE) associated with the loss function of the adjusted model also underwent changes (as shown in Fig.2), and the weight coefficients of the features in the model also changed accordingly (as shown in Fig.3).

**Fig 2.**
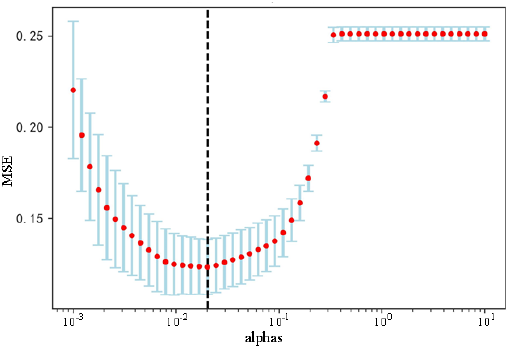
Curve of loss function value with parameter variation

**Fig 3.**
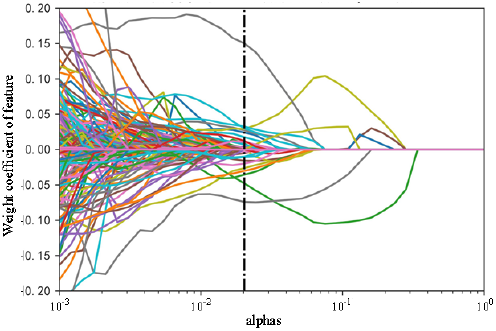
Curve of weight coefficients of feature with parameter variation

The black vertical line in the figure represents the optimal alphas=0.0202, at which point only 28 features with non-zero coefficients remain, namely the dominant features.

When the MSE is at its minimum value, the hyperparameter alphas=0.0202. At this point, the weight coefficients of 729 features in the model become 0. These features with weight coefficients of zero are non-important features and can be removed. The remaining 28 features have non-zero weight coefficients, making them the dominant features. These filtered dominant features and their weight coefficient values are shown in Fig.4.

**Fig 4.**
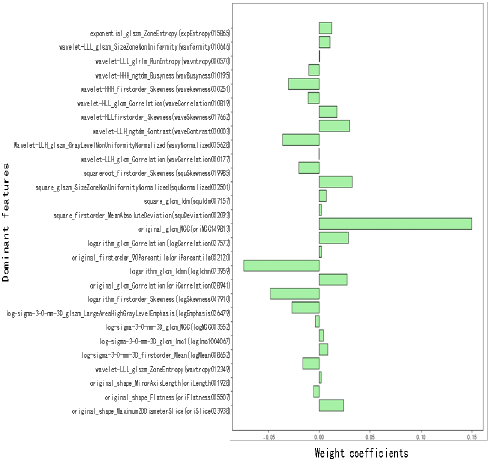
Dominant features along with their weight coefficients

##### Feature validation

Implementing Rstudio 4.4. For our analysis, single-factor logistic regression models were developed to investigate the association between an individual independent variable (often referred to as a feature variable) and the outcome variable (outcome). The analysis of the model’s summary reveals that all 28 of the feature variables possess a significant impact on the classification results (*P <* 0.05).

#### 3.2.2 Development and assessment of radiomics prediction model categorized into four classes

Due to the significant disparity in sample quantities among the four categories of breast density, In the process of classification, the imblearn package’s SMOTE function was utilized to enhance the dataset through up-sampling techniques. Once the processing is completed, all categories show an equal distribution of data.

##### 3.2.2.1 Xgboost model was utilized for classification purposes

Parameter tuning of the Xgboost algorithm was performed. The best hyperparameters were found through 823, 543 iterations of random parameter search. The model based on Xgboost was trained using the dataset for the training phase. was utilized for the classification predictions on the test dataset. The prediction accuracies for classes 0 to 3 were 0.91, 0.87, 0.77, 0.98. The sensitivities for classes 0 to 3 were 0.95, 0.67, 0.85, 0.98. The F1-scores for classes 0 to 3 were 0.93, 0.72, 0.81, 0.98. The total accuracy ACC=0.86 (the overall accuracies for the classes), Cohens Kappa coefficient=0.814. AUC for the four classes and their micro-averaged and macro-averaged values were 1.00, 0.93, 0.93, 0.99, 0.97, 0.96, respectively (as shown in Fig.5).

**Fig 5.**
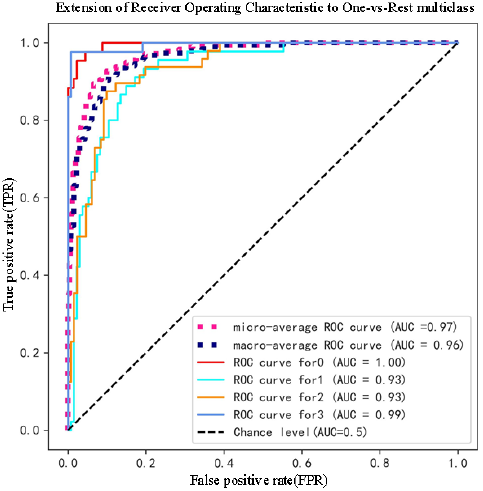
ROC plots for the four classes alongwith their micro-averaged and macro-averaged values for xgboost model

Finally, the calibration curves for the xgboost model performing the four-class classifications of breast density were shown in Fig.6. The decision curves were shown in Fig.7.

**Fig 6.**
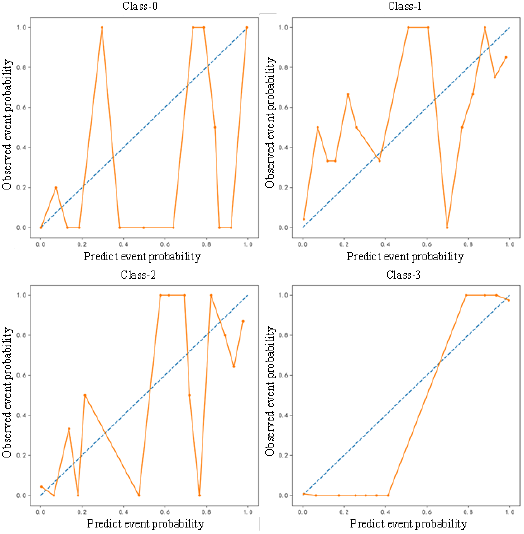
Correction curve for xgboost model of breast density

**Fig 7.**
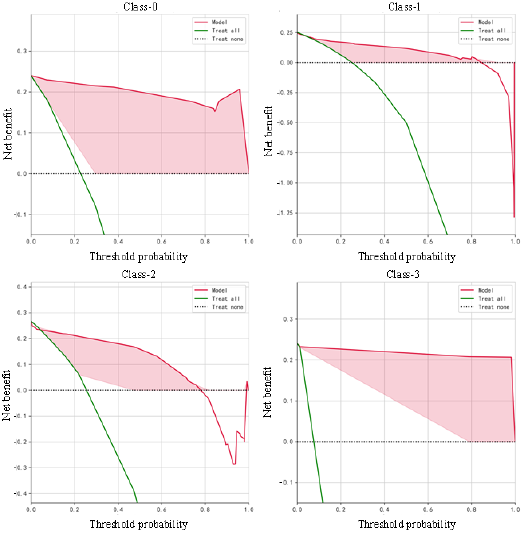
Decision curve for xgboost model of breast density

##### 3.2.2.2 Comparison of four four-class prediction models

Using the same parameter tuning method described above, four four-class prediction models constructed after parameter optimization were as follows: One Vs Rest approach employing Logistic Regression(Kernel), Gradient Boosting Classifier, Random Forest Classifier, and xgboost Classifier. The models fitted with the training set data to the same test set data for classification prediction, and the precisions for each of the class (0-3) predicted by each of the four models were as follows: 0.00, 0.22, 0.00, 0.00; 0.84, 0.79, 0.82, 0.88; 0.87, 0.79, 0.82, 0.90; and 0.91, 0.87, 0.77, 0.98. The sensitivities were 0.00, 1.00, 0.00, 0.00; 0.98, 0.65, 0.72, 0.96; 1.00, 0.68, 0.72, 0.98; 0.95, 0.67, 0.85, 0.98. F1-scores were 0.00, 0.37, 0.00, 0.00; 0.90, 0.71, 0.77, 0.92; 0.93, 0.73, 0.77, 0.94; 0.93, 0.72, 0.81, 0.98. Total accuracy ACC (the overall accuracy for each class) was 0.22, 0.84, 0.85, 0.87. Cohens kappa coefficient was 0.043, 0.783, 0.805, 0.828, respectively.

The areas under the ROC curves (e.g., Fig.8) for each of the four classes and their micro-averaged and macro-averaged values predicted by each of the four models were as follows: 0.73, 0.57, 0.78, 0.54, 0.56, 0.66; 0.99, 0.91, 0.94, 0.99, 0.97, 0.96; 0.99, 0.92, 0.95, 0.99, 0.97, 0.96; 0.99, 0.93, 0.96, 1.00, 0.97, 0.97.

**Fig 8.**
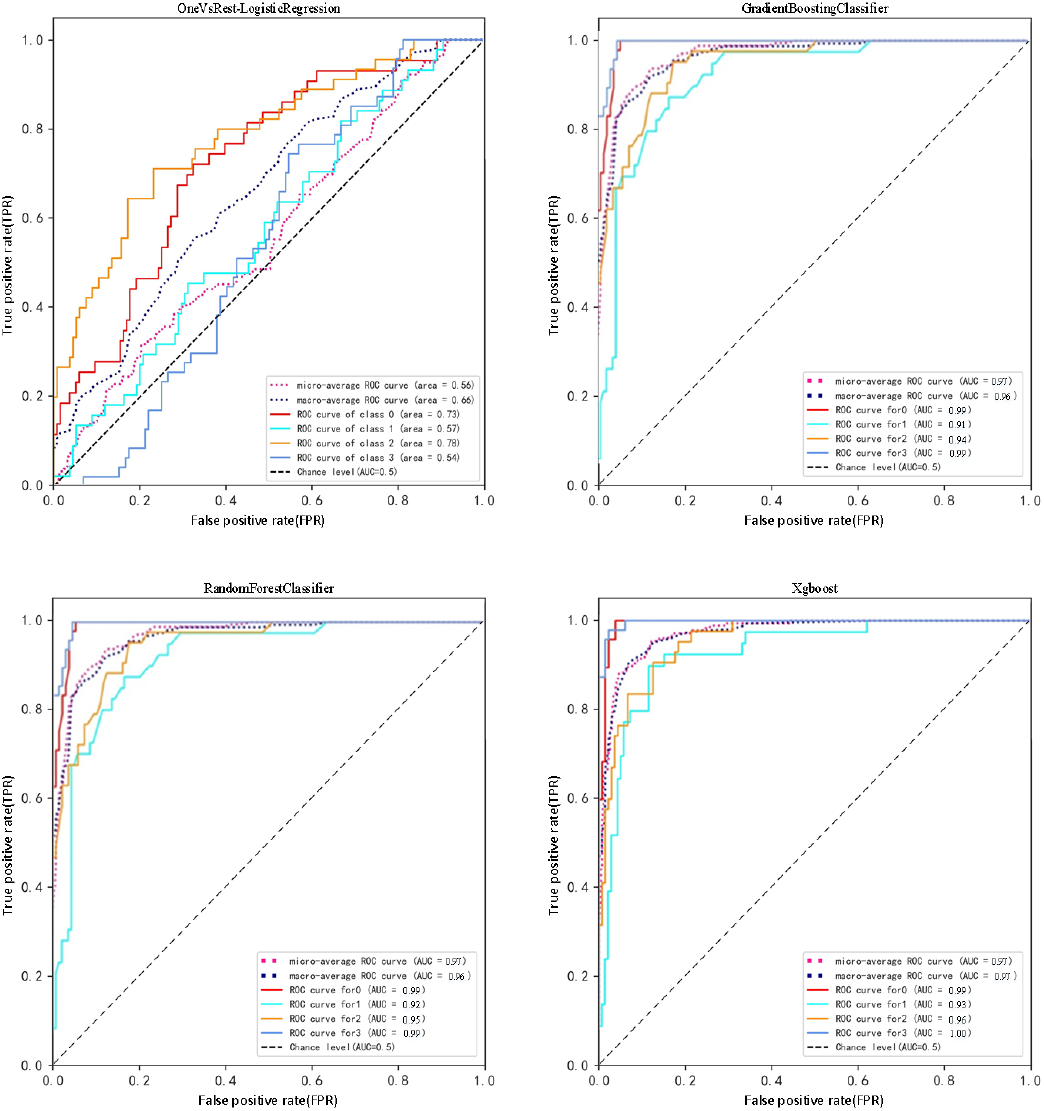
ROC curves obtained when the four models are classified for prediction

The accuracy values were obtained by performing 50 predictions on the test set data using each of the four classification prediction models mentioned above. The average value along with the standard deviation of the performance metrics, including accuracy (ACC) and the 95% confidence interval (CI) pertaining to the accuracy of each model, are outlined as follows: 0.247×0.027[95%CI:0.239-0.255], 0.810×0.029[95%CI:0.802-0.818], 0.822×0.025[95%CI: 0.815-0.829], 0.841×0.028[95%CI:0.823-0.868].

A comprehensive evaluation and comparison of the four models show that: the accuracy of the combined classifier using One Vs Rest multi-classification strategy is very low, less than 50%. The accuracy of the classifiers composed of Gradient Boosting, Random Forest or XGBoost can all reach above 80%. Of these classifiers, the XGBoost model, which has undergone parameter optimization, demonstrates superior predictive performance, and allows for effective utilization in predicting breast density across four distinct categories.

## 4 Discussion

The objective of this research is to automate the categorization of types of breast density by utilizing AI methods. Given the current situation where the volume of chest CT examinations far exceeds that of mammography, this approach can additionally provide breast density information, which is relevant to breast cancer risk, to women through chest CT scans. This, in turn, can help them choose more rational screening options.

Evaluating breast density has demonstrated considerable importance, as it is intricately linked to the risk of developing breast cancer [3, 15–17]. Additional evaluations for women exhibiting greater breast density. can improve the detection rate of breast cancer [4]. Nevertheless, evaluating breast density involves more than just affected by factors such as false positives from ultrasound examinations [18], but also by inter-observer variability. Previous studies have reported poor consistency in breast density assessment based on mammography [19], which can negatively impact the selection of subsequent screening plans.

This study, after blinded evaluation of 393 chest CT and mammography images, demonstrated the consistency of breast density classification between CT and mammography, with CT showing higher inter-observer consistency. This finding is consistent with previous studies using breast-specific 3D imaging techniques, such as digital breast tomosynthesis (DBT) [20], which may indicate that the inter-observer consistency in assessing breast density on 3D images is higher than on 2D images. Therefore, if CT can determine breast density, it would have significant implications for optimizing early breast cancer detection protocols.

The unclear range of breast ROI (Region of Interest) delineating in chest CT has always been a significant challenge for related research. Due to the lack of clearly identifiable boundaries, the issue is more about consistency than accuracy. This research established a distinct scope for the delineation of breast regions of interest based on the anatomical theory of the superficial fascia of the breast [21] and previous experience of imaging review. Although this definition may somewhat enlarge the ROI, it ensures consistency in ROI delineation. The results of this study also confirmed that this delineation criteria ensures the stability and accuracy of AI classification.

Manual ROI delineation is time-consuming, labor-intensive, and subjective. To address these issues, this study developed an automatic segmentation model based on the U-net deep learning algorithm. The model’s Dice coefficient was higher than 0.9, indicating that the segmentation performance is acceptable. Although the ROIs used in the follow-up of this study were manually corrected on the basis of the pre-segmentation by the automatic segmentation model, it is expected that the fully automatic selection of ROIs will be achieved by further improving the accuracy of the segmentation model in future studies.

The research utilized the methodology known as the Least Absolute Shrinkage and Selection Operator (LASSO) algorithm to screen the RFs (Radiomic Features) with ICC (Intraclass Correlation Coefficient) ≥0.75, excluding redundant features. Ultimately, 28 radiomic features closely related to breast density types were identified. Among these, features with larger weight coefficients, such as the maximum correlation coefficient (MCC) from the gray-level co-occurrence matrix (GLCM), describe the texture properties of the image. Features like the inverse variance of the GLCM logarithm describe the uniformity of the image texture, while the logarithm of the first-order skewness describes the skewness of the image gray levels. Overall, the majority of the dominant features belong to the texture properties of the image, which is in accordance with the definition of breast density.

Model training, a variety of four-classification imaging radiomics prediction models were trained, and the AUC values of the optimal model xgboost were 0.97, 0.97, 0.99, 0.93, 0.96, and 1.00 for each classification 0-3 and its micro-averaged and macro-averaged ROC curves, which demonstrated a good evaluative efficacy.

This study established multiple four-classification radiomics predictive models, with the optimal model being xgboost. The area under the ROC curve for the ideal model, xgboost, was recorded at 0.97, 0.97, 0.99, and 0.93 for the class 0-3, respectively, and 0.96 and 1.00 for the micro-average and macro-average, which demonstrated excellent predictive performance.

While the classification models developed in this research demonstrate considerable potential in forecasting, there remain a number of factors that need to be taken into account. To begin with, the research is based on a small dataset and does not have independent external sources for additional verification, which could lead to a selection bias. In addition, the information utilized for this analysis derives from a specific dataset. The construction of the model is limited in scale and may exhibit a skewed distribution of data, which could potentially influence the performance of the model. training. Ultimately, this research concentrated on the classification of breast density using only conventional CT images without additional analysis of breast parenchymal tissue to examine breast density percentages. The current understanding surrounding this matter is insufficient. the utility of this supplementary data. The utilization of artificial intelligence within the domain of breast CT remains in at an initial phase, characterized by technologies and methods that are still developing. Thus, additional investigation is Necessary in the future.

## 5 Conclusion

Based on AI, automatic segmentation of breast ROIs on CT images can be achieved, and the use of appropriate radiomics models can effectively distinguish breast density classification, laying the foundation for achieving precise and personalized breast screening programs.

## Data Availability

All relevant data are within the manuscript and its Supporting Information files.

## Acknowledgment

The study was supported by National Natural Science Foundation of China (No.12226003).

## Availability of data and materialss

The datasets used and analyzed during the current study are available from the corresponding author on reasonable request.

## Author Contributions

Study design: Weipeng Zhou, Huimao Zhang.

Data acquisition: Weipeng Zhou, Qi Yang.

Data analysis and interpretation: Weipeng Zhou, Qi Yang.

Manuscript preparation: Weipeng Zhou.

Critical revision of the manuscript for intellectual content: Weipeng Zhou, Huimao Zhang.

Obtaining financing: Huimao Zhang.

All authors reviewed the manuscript and approved the version to be published.

